# MACHINE LEARNING IMPACT ASSESSMENT OF CLIMATE FACTORS ON DAILY COVID-19 CASES

**DOI:** 10.1101/2022.02.03.22270399

**Authors:** Stephen Afrifa, Essien Felix Ato, Peter Appiahene, Isaac Wiafe, Rose-Mary Owusuaa Mensah Gyening, Michael Opoku

## Abstract

Coronavirus disease 2019 (also known as COVID-19) is a vastly infectious virus instigated by the coronavirus-2, which causes severe acute respiratory illness (SARS-Cov-2). Scientists and researchers are conducting a number of studies to better understand the COVID-19 pandemic’s behavioral nature and spread, and machine learning provides useful tools. We used machine learning techniques to study the effect of climate conditions on daily instances of COVID-19 in this study. The study has three main objectives: first, to investigate the most climatic features that could affect the spread of novel COVID-19 cases; second, to assess the influence of government strategies on COVID-19 using our dataset; third, to do a comparative analysis of two different machine learning models, and develop a model to predict accurate response to the most features on COVID-19 spread. The goal of this research is to assist health-care facilities and governments with planning and decision-making. The study compared random forest and artificial neural network models for analysis. In addition, feature importance among the independent variables (climate variables) were identified with the random forest. The study used publicly available datasets of COVID-19 cases from the World Health Organization and climate variables from National Aeronautics and Space Administration websites respectively. Our results showed that relative humidity and solar had significant impact as a feature of weather variables on COVID-19 recorded cases; and that random forest predicted accurate response to the most climatic features on COVID-19 spread. Based on this, we propose the random forest model to predict COVID-19 cases using weather variables.

## 1 Introduction

Wuhan, Hubei, China, reportedly provided information to the World Health Organization on December 31, 2019 [1] about an undetermined etiology epidemic on coronavirus illness 2019 (often referred to as COVID-19) produced by the coronavirus that causes severe acute respiratory syndrome 2 (SARS-CoV-2). The COVID-19 virus has spread across the globe, affected human activities worldwide, and has become a severe public health problem. The goal of this research is to determine the climate factors in addition to temperature in the daily infection cases instigated by this new coronavirus. COVID-19 infection is similar to the coronavirus that causes severe acute respiratory syndrome (SARS-CoV), could cause serious respiratory disease [2]. According to indirect evidence, 90 percent of non-tropical regions with low temperatures and humidity, COVID-19 cases have been reported up until March 22, 2020, while far fewer instances have been recorded in the tropics [3]. Humidity and Temperature have a big impact on how fast Covid-19 cases proliferate around the world [4]. COVID-19 is transferred and propagated by humans, according to recent investigations [5]. The coronavirus disease of 2019 has clearly spread over the globe and this study aimed to assess the climate impact on daily reported cases for the top ten (10) countries, namely United States, Italy, Spain, Iran, Germany, France, Turkey, United Kingdom, China and Switzerland as of May 2, 2020. Accordingly, it is possible to investigate the COVID-19 transmission as a result of changes in air temperature and relative humidity, and as other researchers found [6]. Many factors can influence virus transmission, including climate (such as humidity and temperature, medical treatment quality, and), population density (Ahmadi et al, 2020). It is hoped that an effective and safe vaccination against the COVID-19 plague, which is instigated by the SARS-CoV-2 coronavirus, would be developed in the future. According to a CDC early release of outcomes among COVID-19 patients in the United States, death was highest in those over 85 years old and usually reduced with age, with no fatalities among those under 19 years old [7]. The main aim of this work is to develop a Machine Learning algorithm model to assess the impact of climate conditions on daily recorded cases of COVID-19 for the topmost ten (10) countries in the world. The study also specifically aims to investigate the most climatic features that could affect the spread of novel COVID-19 cases, to assess the influence of government strategies on COVID-19 using dataset from WHO, and to do a comparative analysis of two different Machine Learning models, and develop a model to predict accurate response to the most climatic features on COVID-19 spread. This study proposes a predictive model that can predict the most climatic feature regarding the emergence of new COVID-19 instances among the top ten (10) countries as of May 2, 2020, using Random Forest and Artificial Neural Network. This will help to assess the impact of climate features in the increasing daily recorded cases of COVID-19 in the world. News reports, publications, preprints, and white papers have received a lot of attention in this study in a directive to control the COVID-19 pandemic in 2020, which is triggered by SARS-CoV-2 (Karapiperis et al., 2020). The ability of the models to predict and compare the climatic conditions will help researchers and health officers to make concrete decisions on the behavioral dynamics of the coronavirus, and give opportunity to policymakers and governments across the globe to make decisions. The study is conducted on the COVID-19 data from the World Health Organization (WHO) website. The target is to predict and compare the most climatic feature on the occurrence of new COVID-19 cases every day in Spain, Italy, the United Kingdom, the United States, Germany, France, China and Switzerland, Turkey, Iran using two (2) Machine Learning models.

## 2 Related Works

We were able to uncover the resulting relevant works, which primarily consist of studies utilizing machine learning models, with the goal of forecasting the impact of meteorological conditions on COVID-19 confirmed cases. For example, Pramanik *et al*. (2020) looked at the impact of weather on the spread of COVID-19 in Russia. In the humid continental zone, temperature seasonality (29.2 0.9 percent) had the greatest impact on COVID-19 transmission., according to the findings of their study. The diurnal temperature range (26.8 0.4 percent) and temperature seasonality (14.6 0.8 percent) had the biggest impacts in the subarctic region. In another related work by Mehmet (2020) the researcher looked at nine places in Turkey to see if there was a link between weather and coronavirus disease 2019 (COVID-19). Dew point (°C), humidity (percent), temperature (°C), and wind speed (mph) were used as weather parameters in the study. The correlation coefficients of Spearman were used to conduct the studies. The population, temperature, and wind speed 14 days ago on the day had the strongest relationships, according to the findings.

Infections with COVID-19 and fatalities in Mexico were projected using machine learning techniques by Gomez *et al*. (2021) in their study. Their study has three main objectives: first, to discover which function in Mexico best responds to the increase in infected people; second, to assess the significance of climatic and mobility characteristics; third, to compare the outcomes of a standard time series statistical model with those of a new machine learning technique. To explain the rise of COVID-19 episodes in Mexico, polynomial, linear, and extended logistic regression models were compared.

Karapiperis *et al*. (2021) used machine learning to show that UV radiation was more significantly connected significantly higher rates of occurrence than mobility, showing that UV is a major Covid-19 seasonality indication, regardless of the epidemic’s beginning circumstances. Hass and Arsanjani (2021)) suggested a data-driven strategy for examining the pandemic’s spatio-temporal patterns on a regional size, such as Europe, and a country scale, such as Denmark, as well as what geographical variables may contribute to its spread. For studying the possible association amid the selected underlying parameters and pandemic spread, they used statistical approaches such as machine learning approaches such as random forest, as well as ordinary least squares and regionally weighted regression.

In a study published by Xie and Zhu (2020) sought to examine whether temperature plays a role in the infection produced by this new coronavirus. Between January 23, 2020, and February 29, 2020, confirmed cases on a daily basis, as well as weather parameters were gathered in 122 cities, and the nonlinear relationship between mean temperature and COVID-19 confirmed cases was investigated using a generalized additive model (GAM). Piecewise linear regression was also used to find the link. Furthermore in New York City, USA, Farhan *et al*. (2020) looked into the link between COVID-19 and climatic indices. Lowest temperature, maximum temperature, average temperature, rainfall, average humidity, air quality and wind speed are among the climate indicators studied. For data analysis, Kendall and Spearman rank correlation tests were performed. The minimum temperature, average temperature and air quality were discovered to be strongly connected to the COVID-19 pandemic in their research. In Jakarta, Indonesia, Tosepu *et al*. (2020) explored the association between weather and the covid-19 pandemic in another study. The data was analyzed using the Spearman-rank correlation test. Only the average temperature (in degrees Celsius) was substantially linked with the covid-19 pandemic (r = 0.392; p b.01) among the weather variables.

Moreover, Ma *et al*., (2020) in a separate investigation, researchers found a link between coronavirus disease (COVID-19) death and weather conditions. They collected day-to-day COVID-19 death numbers, as well as meteorological and air pollutant data, from January 20 through February 29, 2020, in Wuhan, China. Using the generalized additive model, the effect of temperature, humidity, and diurnal temperature range on COVID-19 daily mortality was examined. Temperature variation and humidity, according to the model, may be key factors determining COVID-19 death. In their study, Liu *et al*. (2020) looked at the correlations between the novel coronavirus disease 2019 (COVID-19) case count and climatic parameters in 30 Chinese province capital cities. Using non-linear regression, they looked at the relationships between the verified COVID-19 case counts, MSI and climatic parameters. Then there were 17 cities with over 50 verified cases, they did a two-stage study. Ahmadi *et al*. (2020) investigated the effect of climatic conditions on the spread of COVID -19 in Iran in a related study. The number of patients infected with COVID-19, population density, and intra-provincial mobility are all factors to consider, average temperature, average precipitation, humidity, wind speed, and average solar radiation from infection days until the end of the study period were the primary characteristics.

## 3 Methodology

### 3.1 Data Collection

In this study, datasets of weather features and COVID-19 recorded cases were used to train the models developed in this work.

#### 3.1.1 The COVID-19 Datasets

COVID-19 cases (as a dependent variable) from Spain, Italy, the United Kingdom, the United States, Germany, Turkey, Iran, France, China, and Switzerland were acquired from the World Health Organization’s official website (https://covid19.who.int). As of January 1, 2020 to May 2, 2020 when the data was collected for this study, the understudied countries all together recorded 2,330,216 cases, with the United States recorded the highest with Switzerland having the least recorded cases. Table 1 shows the results.

**Table 1:**
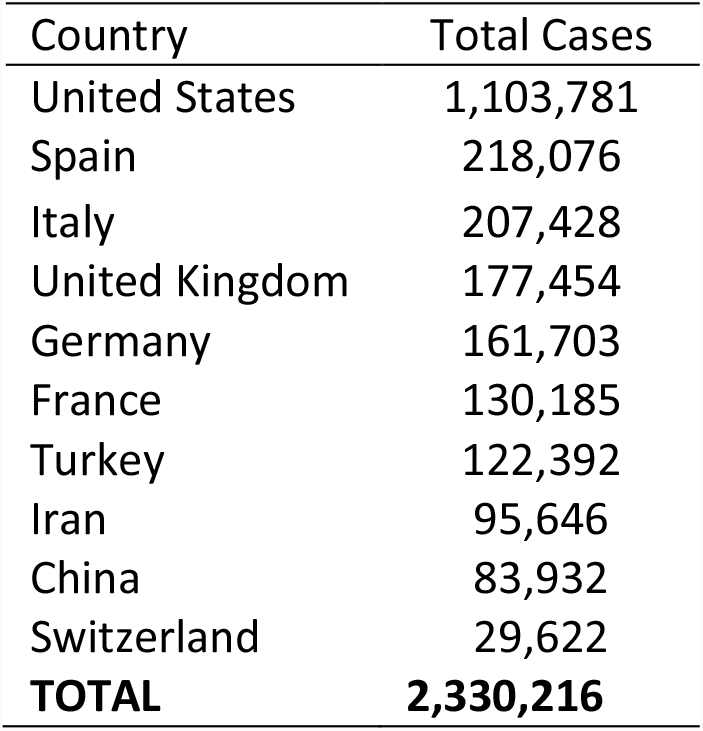
Total recorded cases of COVID-19 (January 1, 2020 to May 2, 2020)

**Figure 1** depicts the situation of new COVID-19 cases in the understudied nations from January 1, 2020 to May 2, 2020.

**FIgure 1:**
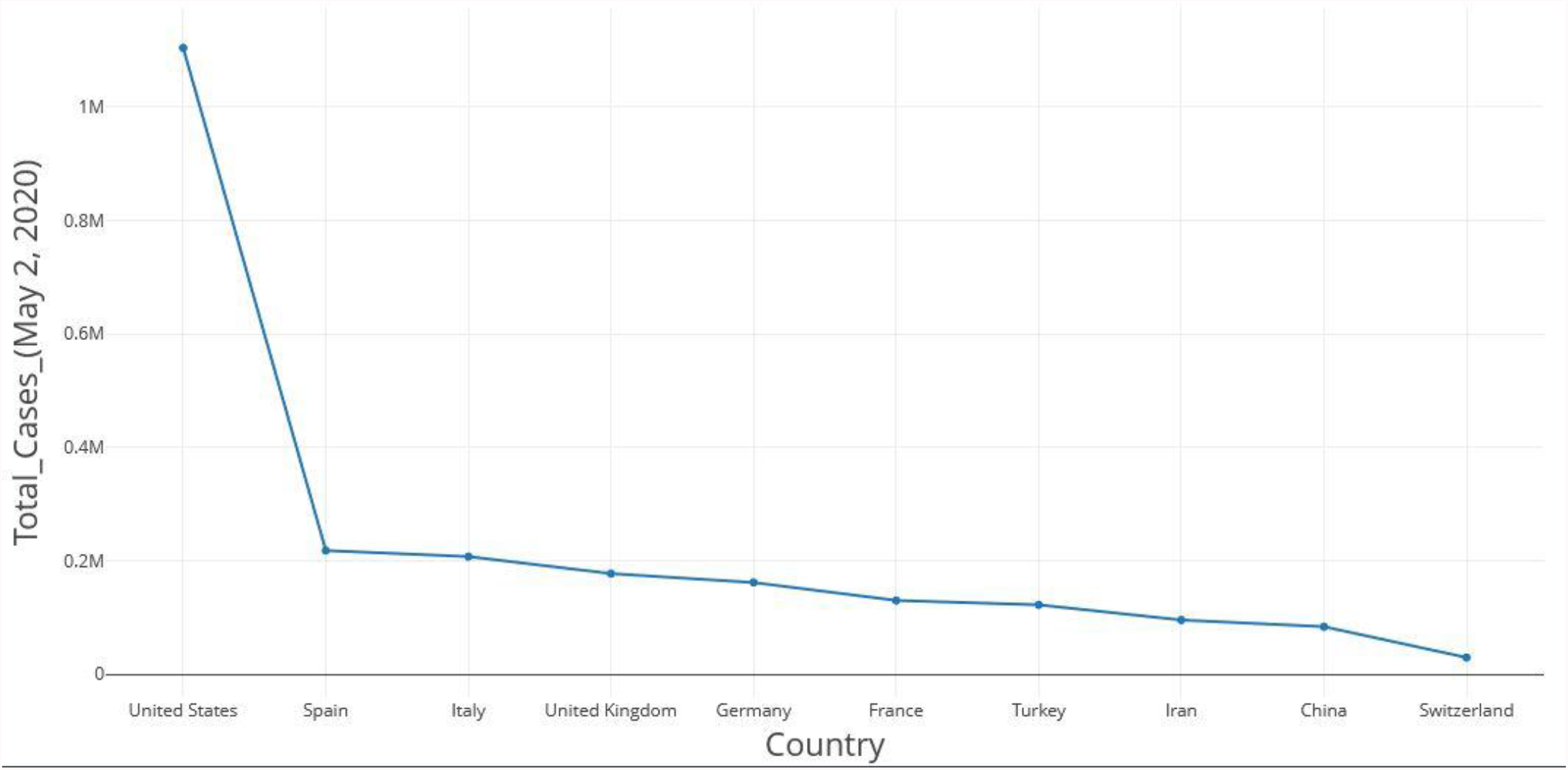
New COVID-19 cases from January 1, 2020 to May 2, 2020

The daily recorded cases of the COVID-19 dataset were divided and grouped in ranges by the researchers as shown in table 2 below;

**Table 2:** Dependent (COVID-19 cases) Variables for our study

| Dependent Variables<br>(Levels) | Range (Recorded<br>COVID-19 Cases) |
| --- | --- |
| Level1 | 0 – 1999 |
| Level2 | 2000 – 4999 |
| Level3 | 5000 – 9999 |
| Level4 | 10000 – 29999 |
| Level5 | 30000+ |

#### 3.1.2 The Weather Dataset

The National Aeronautics and Space Administration (NASA) webpage (https://power.larc.nasa.gov), provided the information on climate data. The information retrieved of weather were based on 9 conditions from the understudied countries as of January 1, 2020 to May 2, 2020. The extracted features for this study are the following: precipitation, relative humidity at 2meters, maximum and minimum temperature at 2meters, maximum wind at 10meters, wind speed at 50meter, and solar (isolation clearness index, all sky insolation incidence on a horizontal surface, downward thermal infrared (longwave) radiative flux) as shown in table 3 below;

**Table 3:**
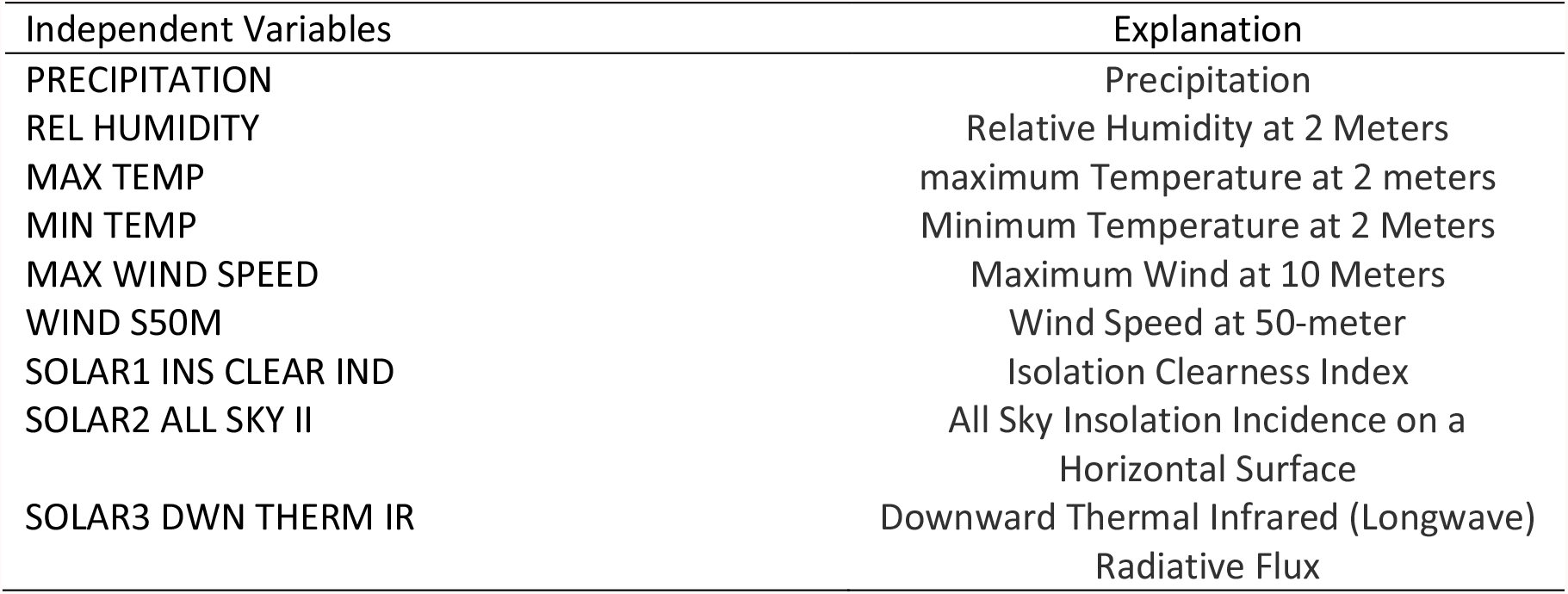
Predictor Variables for our study

| Independent Variables | Explanation |
| --- | --- |
| PRECIPITATION | Precipitation |
| REL HUMIDITY | Relative Humidity at 2 Meters |
| MAX TEMP | maximum Temperature at 2 meters |
| MIN TEMP | Minimum Temperature at 2 Meters |
| MAX WIND SPEED | Maximum Wind at 10 Meters |
| WIND S50M | Wind Speed at 50-meter |
| SOLAR1 INS CLEAR IND | Isolation Clearness Index |
| SOLAR2 ALL SKY II | All Sky Insolation Incidence on a<br>Horizontal Surface |
| SOLAR3 DWN THERM IR | Downward Thermal Infrared (Longwave)<br>Radiative Flux |

## 3.2 Methods

Using machine learning, this study investigates the impact of climate conditions on COVID-19 instances on a daily basis. The data analysis and machine learning models were created using the R programming language, which was run on the open-source R Studio platform. Two machine learning models were used in our approach, thus, Random Forest (RF) and Artificial Neural Networks (ANN) on classification for this analysis based on the main and specific objectives. The target for the model is the daily cases, which is divided and grouped between level 1 to 5, thus, to minimize the training time and increase the model’s performance. The diagram below shows the proposed model.

### 3.2.1 Random Forest (RF) Algorithm

Breiman in 2001 and cited by (Appiahene et al., 2020) said the Random Forest (RF) algorithm is used for classification and prediction that employs an ensemble of classification trees. Random Forest is a learning method that entails the construction of many decision trees. The final decision is made by the random forest, which is based on the majority of the trees. Each tree in the random forest predicts a class, and the class with the most votes become our model’s forecast. The model can generate a number to show the spatial relationships, decision-making trees are used between COVID-19 cases and the environmental parameters studied in this study. A random forest model is more capable of modeling and forecasting according to (Kane et al., 2014) and understood better with the steps below;

- **Step 1** − Begin by randomly choosing samples from a dataset.
- **Step 2** − Following that, for each sample, this algorithm will generate a decision tree. After that, each decision tree’s predicted result will be obtained.
- **Step 3** − During this step, you will be able to vote on each projected outcome.
- **Step 4** − Finally, as the final prediction result, choose the prediction result with the most votes.

The Random Forest algorithm;

~~~
***procedure*** *RANDOM FOREST
         for* ***1*** *to T* ***do***
                    *Draw n points D*_*i*_ *with standby from D
                    Build full decision or regression tree on D*_*i*_
                               *But: Each split only reflect k features, picked homogenously at random
                                       new features for every split
                    Trim tree to minimalize out-of-bag error*
        ***end for
        Average*** *all T trees*
***end procedure***
~~~

A diagram of Random Forest Model is represented below;

#### 3.2.2 The Artificial Neural Network

According to a citation by (Appiahene et al., 2020), an artificial neural network (ANN) is a sort of Artificial Intelligence (AI) technique that mimics the behavior of a human brain. Artificial Neural Networks (ANN) are a data processing model based on how the biological nervous system, such as the brain, processes information. The ANN architecture is explained below;

- The first is the Information layer, which contains the units that take input from the outside world and analyze, identify, or otherwise process it.
- The next layer is the Output layer, which comprises units that respond to data on how it learned any mission.
- The job of the hidden layer is to transform the input into something that the output unit can use. The Hidden layer is in-between input layer and output layer. Artificial Neurons are the units in the Input Layer.

The Artificial Neural Network algorithm;

*For each output note compute*

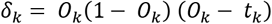

*For each hidden node calculate*

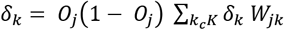

*Bring up-to-date the weights and biases as follows*

*Given*

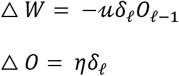

*apply*

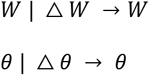

A diagram for Artificial Neural Network below;

## 4. Results and Discussion

This section summarizes our findings and contributions made to academia. Our findings on climatic effect on daily cases of COVID-19 showed that some weather features (predictors) are ranked higher which much emphasis placed accordingly in this study. The figure 5 shows the top-ranking climatic variables used to build the model.

**Figure 2:**
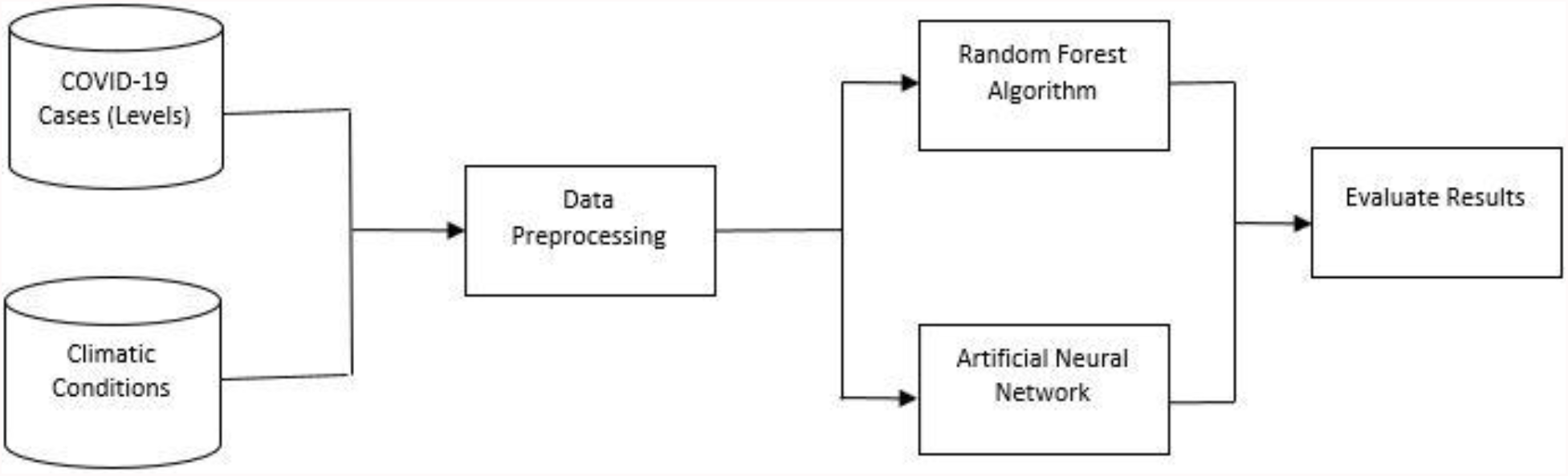
The proposed model (Source: Author’s own)

**Figure 3:**
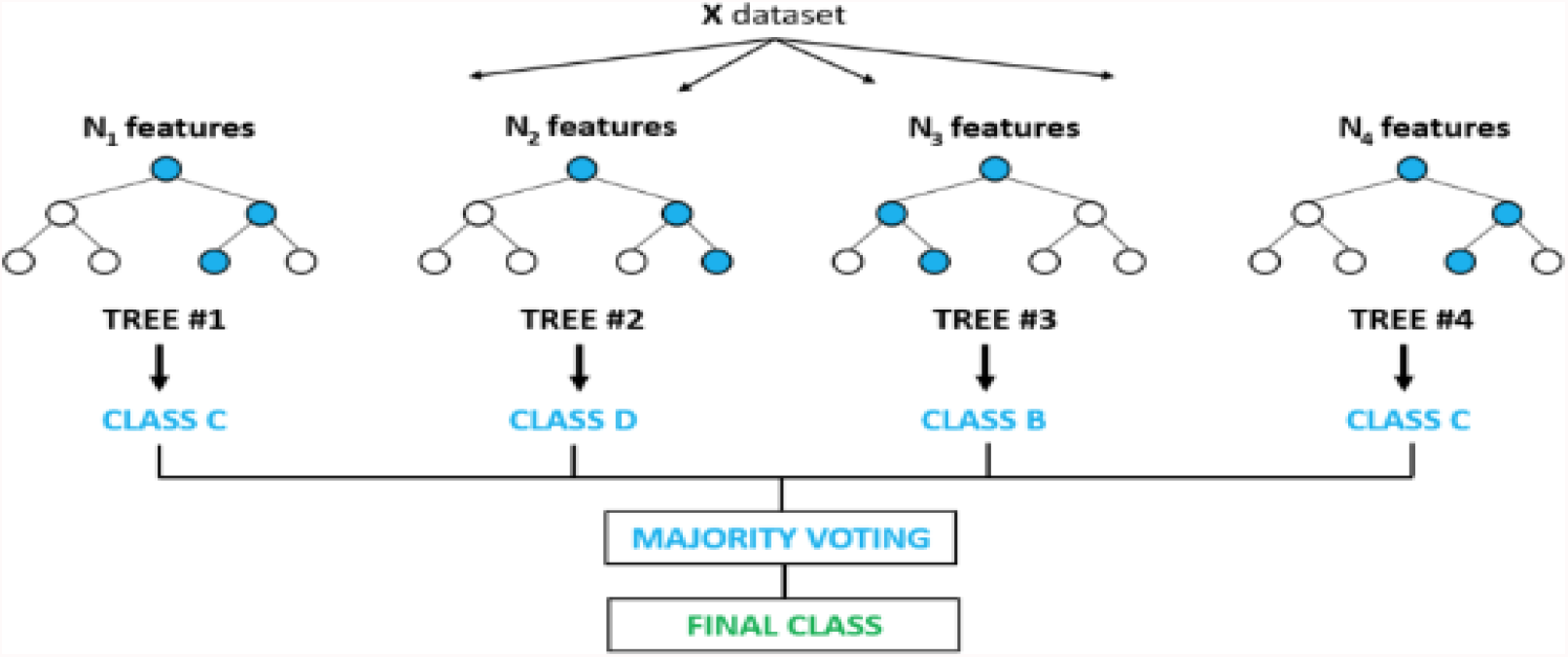
Random Forest Example (Source: Kaitlin et al., 2018)

**Figure 4:**
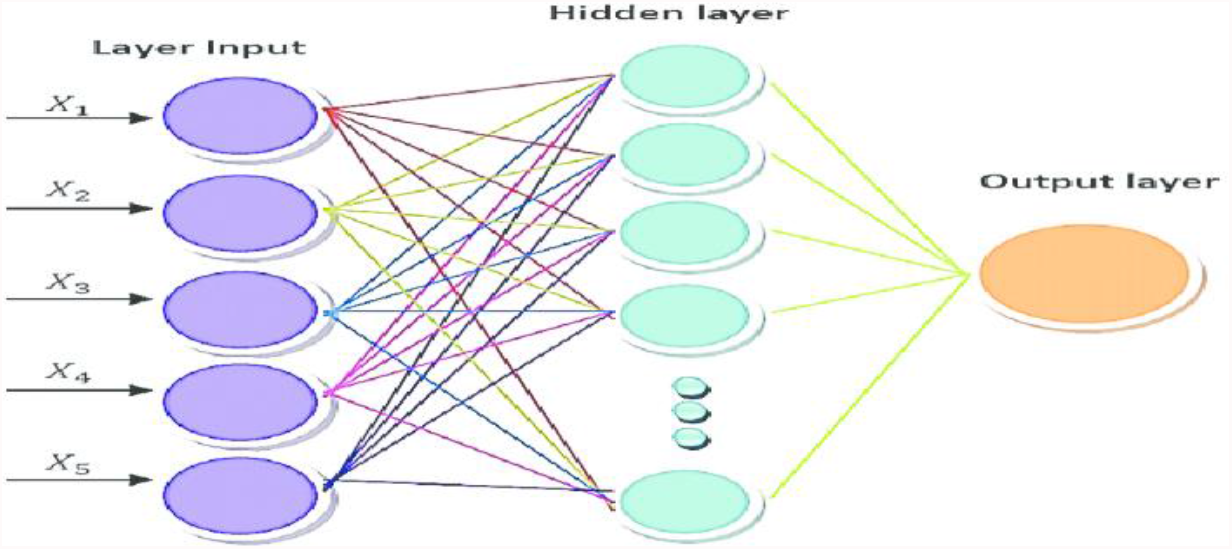
Diagram of ANN Model (Source: Khademi et al., 2015)

**Figure 5:**
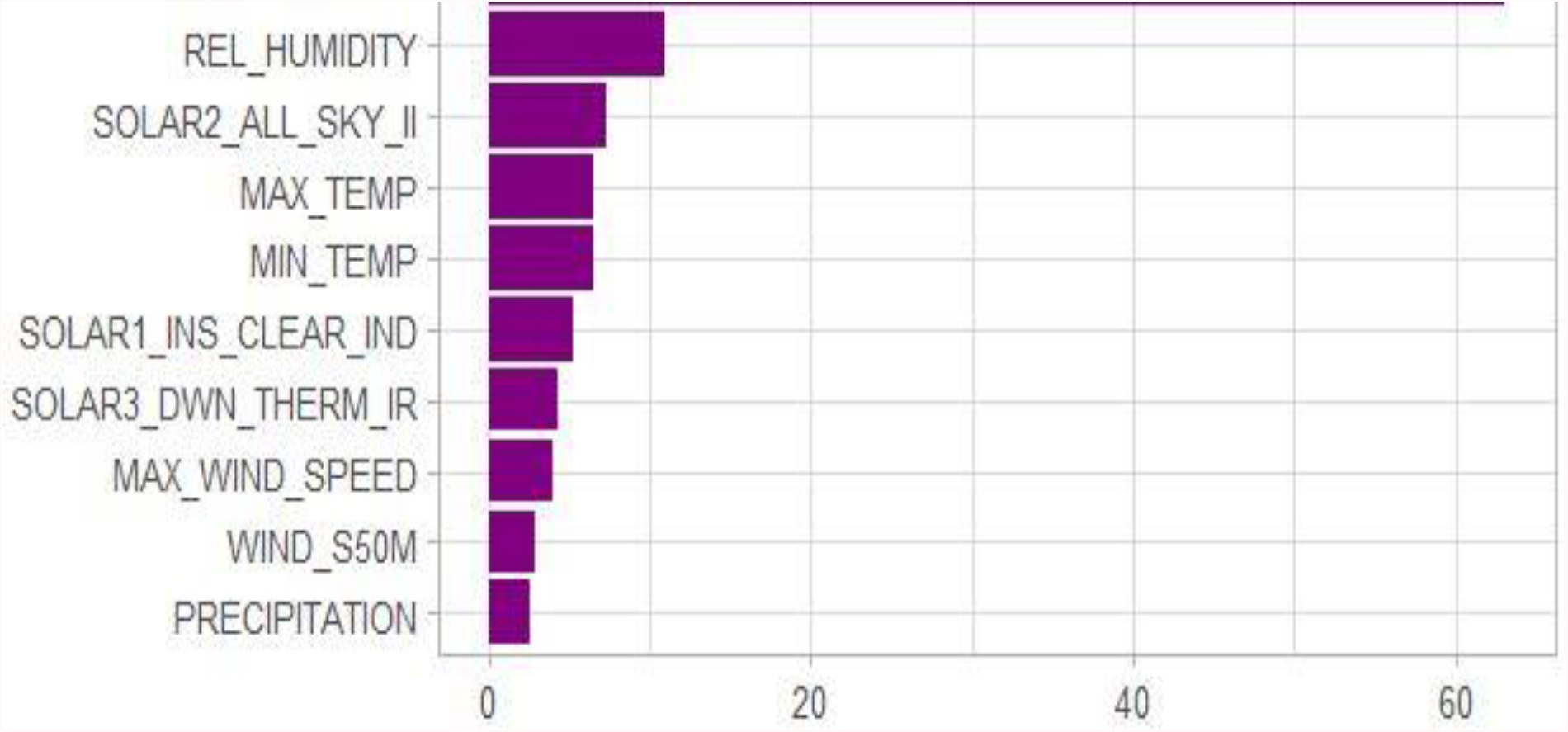
Top ranking predictor (climatic) variables (Source: Author’s construct).

In the table 4 we report the models that were trained on the weather features and COVID-19 data with their evaluation metrics.

**Table 4:** Models’ evaluation

| Table 4: Models' evaluation |  |  |  |
| --- | --- | --- | --- |
| Model | Accuracy (%) | RMSE | MAE |
| Random Forest (RF) | 99.71 | 0.62 | 0.30 |
| Artificial Neural Network (ANN) | 68.70 | 0.57 | 0.21 |

All features in the weather dataset were used in the training of models. The daily cases were grouped in ranges, thus, to reduce training time and increase models’ performance. The results showed that Random Forest (RF) was the best performing model. The extracted features were ranked according to influence on the results. Both developed models predicted that relative humidity is the highest-impact weather parameter followed by solar. The developed model of Random Forest achieved the highest accuracy. Moreover, it can be observed that both models performed better the RF has a better performance in relation to our main and specific objectives in this study.

Our findings are consistent with those of Pramanik *et al*., (2020) about the important role relative humidity impacts on coronavirus. The trend of daily cases is shown in Figure 6, and relative humidity is shown in Figure 7 between the ten countries considered in this paper. It is evident that the number of infections in the countries were at its peak between February 2020 and April 2020, but the United States recorded more in this study.

**Figure 6:**
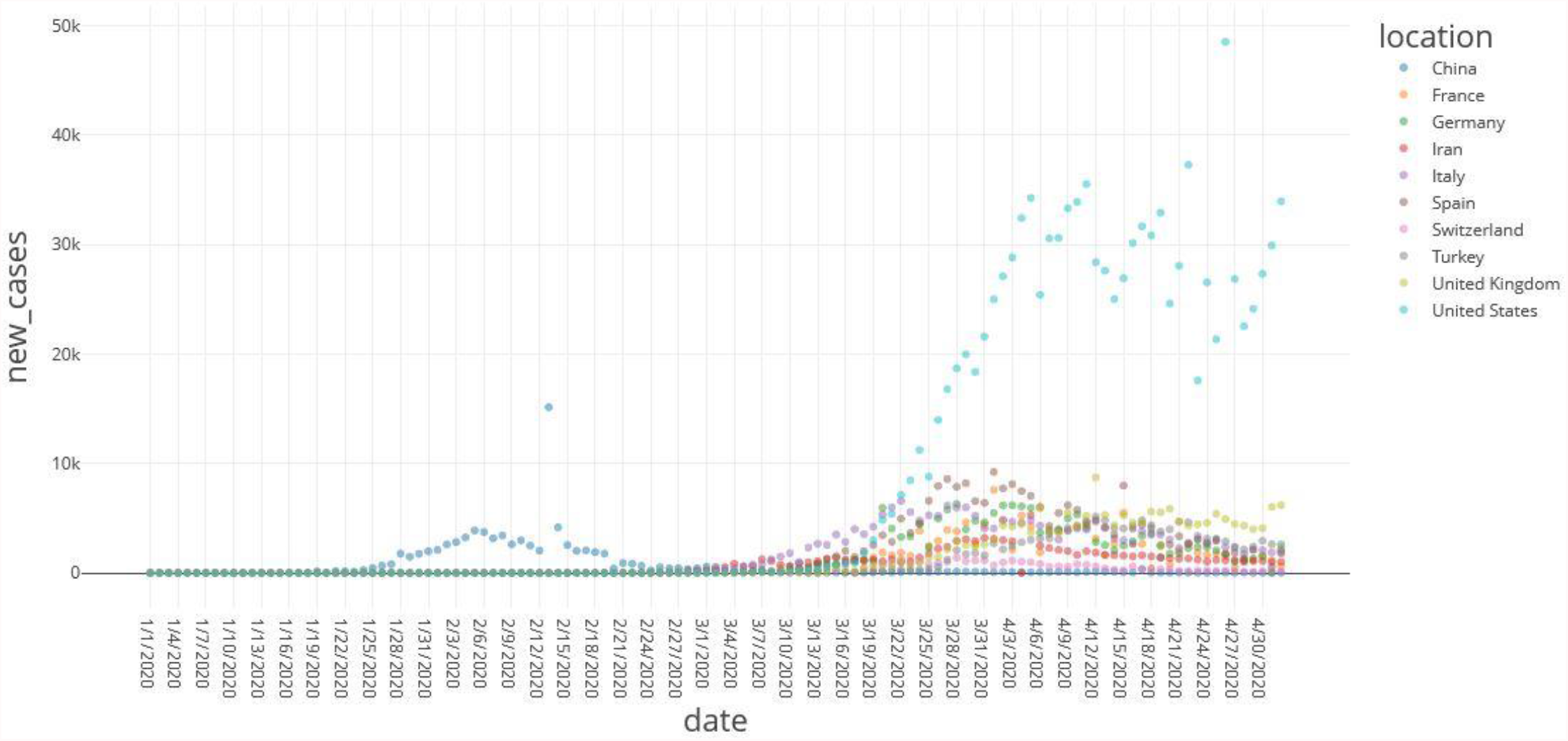
COVID-19 cases as confirmed in ten countries

**Figure 7:**
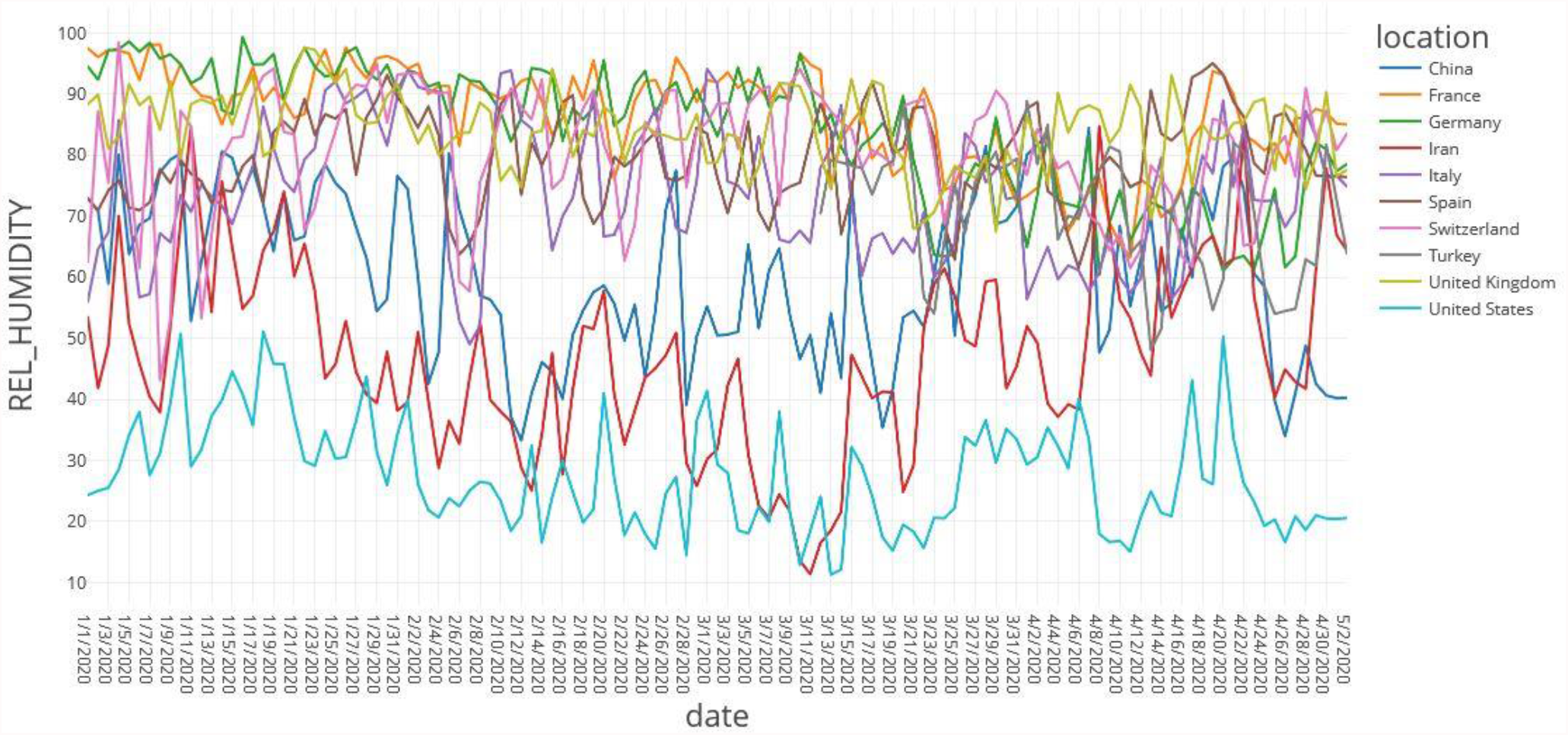
Relative humidity in ten countries

Figure 8 below represents the map of the countries which were taken into consideration in this study as of January 1, 2020 to May 2, 2020.

**Figure 4:**
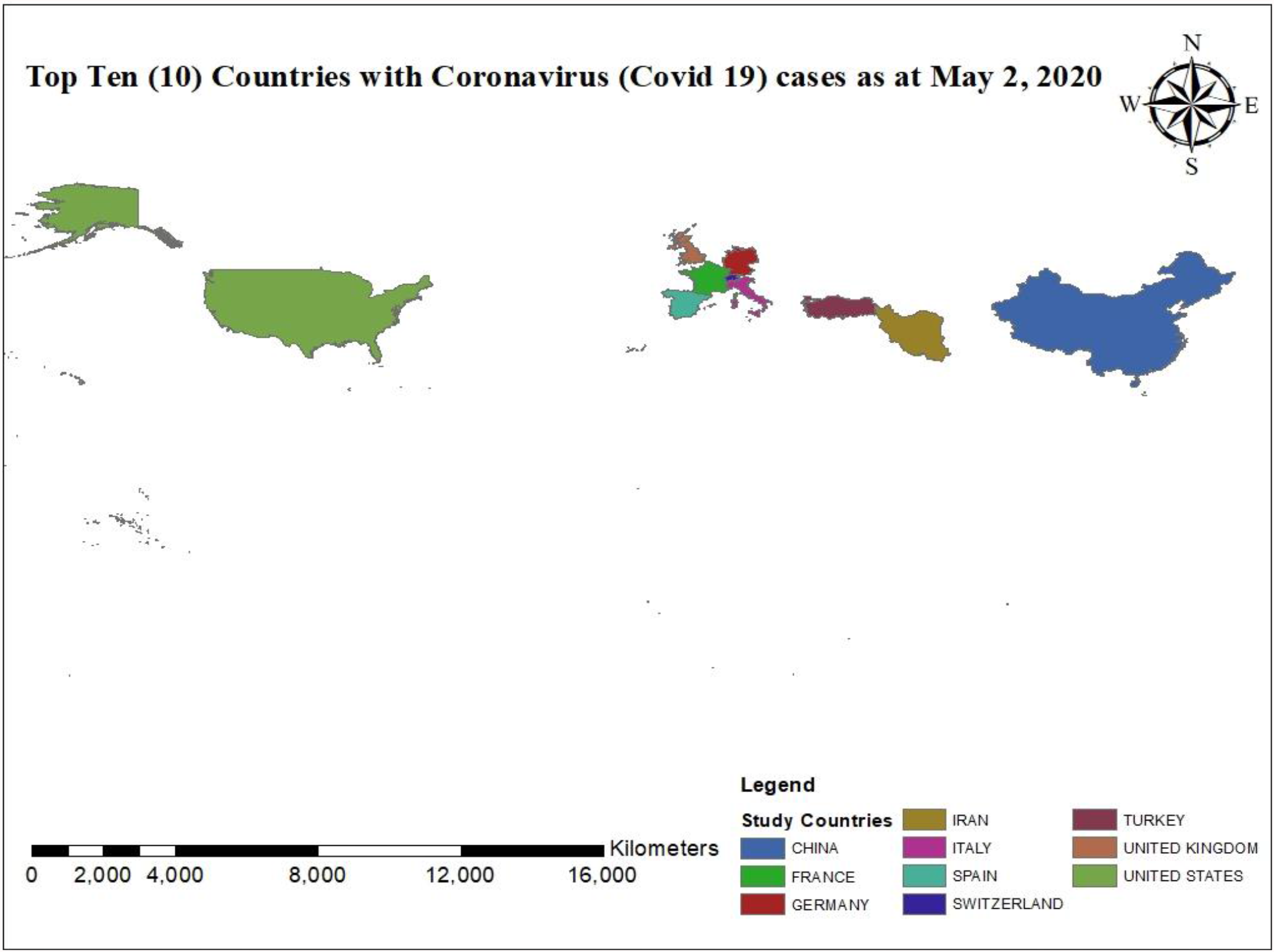
Top ten (10) countries understudied (Source: Author’s construct)

## 5.1 Conclusion

Today, the world is striving for technology and its impact is very immense. Machine Learning Algorithms are making things easier, this is because machines learn from precedence and estimate future events. The researchers employed machine learning techniques to study the effect of climate on COVID-19 cases on a daily basis. The study also aimed at investigating the most climatic feature; analyzing the influence of government measures on COVID-19; preventing the proliferation of additional COVID-19 instances using our dataset; and comparatively analyzing the different machine learning models. Moreover, and developed a model to predict accurate response to the most climatic features on COVID-19 infection. This study used the Random Forest and Artificial Neural Network (backpropagation algorithm) models in building our model to estimate the effect of climatological conditions on COVID-19; from January 1, 2020 to May 2, 2020; for the top ten (10) countries. The countries understudied are Spain, Italy, Turkey, Iran, the United Kingdom, the United States, Germany, France, China, and Switzerland respectively. Our models were based on the classification aspect of Random Forest and Artificial Neural Network. Our datasets were collected from the National Aeronautics and Space Administration (NASA) and the World Health Organization (WHO) websites for climatic variables and COVID-19 cases respectively. Precipitation, maximum temperature at 2 meters, relative humidity, all-sky insolation incidence on a horizontal surface, at 2 meters, downward thermal infrared (Longwave) radiative flux, isolation clearness index, wind speed at 50 meters, minimum temperature at 2 meters, and maximum wind at 10 meters were the independent variables. The dependent variable was the level attribute based on recorded cases of COVID-19 divided into ranges, which are, Level1, Level2, Level3, Level4, and Level5. The results predicted that, relative humidity followed by solar have the greatest influence on recorded cases of COVID-19 in the understudied countries. From our study, Random Forest showed a higher accuracy followed by Artificial Neural Network using our dataset. Based on our analysis and evaluation metrics, it is predicted that Random Forest algorithm can give precise forecasting when it comes to investigating the effect of weather on COVID-19 cases. Nevertheless, our data sample consisted of ten selected countries, but the model can be used to assess climate features of novel cases of COVID-19 across various continents.

### 5.2 Recommendations and Future Works

For future instructions, we recommend that the dataset be increased than what was used in this project, as it can help improve the accuracies more than what was obtained in this project. Moreover, at least twenty (20) countries or more can be as compared with the same feature attributes from our dataset. Furthermore, we recommend that the AI community, the medical community, developers, and policymakers collaborate and share data on a multidisciplinary and multi-stakeholder basis, both nationally and internationally, identify relevant data and open datasets, share tools, to formulate the problem and train models to effectively combat this pandemic. This would help scientists to perform as many Machine Learning algorithm models to know the behavior, the spread, and mitigate ways in the confrontation of the virus. There are lots of emerging COVID-19 variants which makes it difficult to combat the disease easily presently – our dataset can be used to assess the impact on these new/possible variants in the future.

## Data Availability

The data used in this study is available online via 

https://figshare.com/articles/dataset/climatic_features_and_COVID19_csv/19116071

## Data Availability

The CLIMATIC_FEATURE_AND_COVID19 data used to support the findings of this study have been deposited in the FIGSHARE repository with the following link climatic_features_and_COVID19.csv (figshare.com)

## Conflict of Interest

The editors declare that they have no conflicts of interest regarding the publication of this special issue.

## Funding Statement

This is a self-funded project and as a means to contribute to academia.

## Acknowledgements

We are grateful to the World Health Organization (WHO) and National Aeronautics Space Agency (NASA) for making the datasets available on their websites.

## Notes

### Competing Interest Statement

The authors have declared no competing interest.

### Funding Statement

This study is a self funded project

### Author Declarations

The study used openly available data which can be obtained from the link below; https://figshare.com/articles/dataset/climatic_features_and_COVID19_csv/19116071

## References

[1] J. Liu et al., “Impact of meteorological factors on the COVID-19 transmission : A multicity study in China,” Sci. Total Environ., vol. 726, p. 138513, 2020.

[2] J. Xie and Y. Zhu, “Association between ambient temperature and COVID-19 infection in 122 cities from China,” Sci. Total Environ., vol. 724, p. 138201, 2020.

[3] J. Wang, K. Tang, K. Feng, and W. Lv, “High Temperature and High Humidity Reduce the Transmission of COVID-19,” 2020.

[4] G. F. Ficetola and D. Rubolini, “Climate Affects Global Patterns Of Covid-19 Early Outbreak Dynamics,” pp. 1–24, 2020.

[5] Ş. Mehmet, “Impact of weather on COVID-19 pandemic in Turkey,” vol. 728, 2020.

[6] N. M. T. Jebril, “Predict the transmission of COVID-19 under the effect of air temperature and relative humidity over the year in Baghdad, Iraq,” 2020.

[7] M. Ahmadi, A. Sharifi, S. Dorosti, S. J. Ghoushchi, and N. Ghanbari, “Investigation of Effective Climatology Parameters on COVID-19 Outbreak in Iran,” Sci. Total Environ., p. 138705, 2020.

[8] C. Karapiperis, P. Kouklis, S. Papastratos, A. Chasapi, C. A. Ouzounis, and C. Process, “Assessment for the seasonality of Covid-19 should focus on ultraviolet radiation and not ‘warmer days,’” pp. 19–20, 2020.

[9] M. Pramanik, P. Udmale, P. Bisht, K. Chowdhury, S. Szabo, and I. Pal, “Climatic factors influence the spread of COVID-19 in Russia,” Int. J. Environ. Health Res., vol. 00, no. 00, pp. 1–15, 2020.

[10] D. A. Gomez, C. Ramon, E. D. Ramos, F. J. Cantu, and O. Hector, “Data Analysis and Forecasting of the COVID - 19 Spread : A Comparison of Recurrent Neural Networks and Time Series Models,” Cognit. Comput., no. 0123456789, 2021.

[11] C. Karapiperis et al., “A Strong Seasonality Pattern for Covid-19 Incidence Rates Modulated by UV Radiation Levels,” pp. 1–17, 2021.

[12] F. S. Hass and J. J. Arsanjani, “The Geography of the Covid-19 Pandemic : A Data-Driven Approach to Exploring Geographical Driving Forces,” 2021.

[13] M. Farhan, B. Ma, B. Komal, M. Adnan, D. Tan, and M. Bashir, “Correlation between climate indicators and COVID-19 pandemic in New,” Sci. Total Environ., vol. 728, p. 138835, 2020.

[14] R. Tosepu et al., “Correlation between weather and Covid-19 pandemic in Jakarta, Indonesia,” vol. 725, 2020.

[15] Y. Ma et al., “Effects of temperature variation and humidity on the mortality of COVID-19 in Wuhan,” 2020.

[16] P. Appiahene, Y. M. Missah, and U. Najim, “Predicting Bank Operational Efficiency Using Machine Learning Algorithm: Comparative Study of Decision Tree, Random Forest, and Neural Networks,” Adv. Fuzzy Syst., vol. 2020, no. July, pp. 1–12, 2020.

[17] M. J. Kane, N. Price, M. Scotch, and P. Rabinowitz, “Comparison of ARIMA and Random Forest time series models for prediction of avian influenza H5N1 outbreaks,” BMC Bioinformatics, vol. 15, no. 1, 2014.

[18] ;Kaitlin T.; Smith, and B. Sadler, “Random Forest vs Logistic Regression: Binary Classification for Heterogeneous Datasets,” SMU Data Sci. Rev., vol. 1, no. 3, p. 9, 2018.

[19] F. Khademi, M. Akbari, and S. M. Jamal, “Prediction of Compressive Strength of Concrete by Data-Driven Models,” i-manager’s J. Civ. Eng., vol. 5, no. 2, pp. 16–23, 2015.

